# Identification of carcinogenesis and tumor progression processes in pancreatic ductal adenocarcinoma using high-throughput proteomics

**DOI:** 10.1101/2022.03.31.22273227

**Authors:** Lucía Trilla-Fuertes, Angelo Gámez-Pozo, María Isabel Lumbreras-Herrera, Rocío López-Vacas, Victoria Heredia-Soto, Ismael Ghanem, Elena López-Camacho, Andrea Zapater-Moros, María Miguel, Eva M Peña-Burgos, Elena Palacios, Marta de Uribe, Laura Guerra, Antje Dittmann, Marta Mendiola, Juan Ángel Fresno Vara, Jaime Feliu

## Abstract

Pancreatic ductal adenocarcinoma (PDAC) is an aggressive disease with an overall 5 year-survival rate of just 5%. A better understanding of the carcinogenesis processes and the mechanisms of progression of PDAC is mandatory.

Fifty-two PDAC patients treated with surgery and adjuvant therapy, with available primary tumor, normal tissue, preneoplastic lesions (PanIN), and/or lymph node metastases, were selected for the study. Proteins were extracted from small punches and analyzed by LC-MS/MS using data-independent acquisition. Proteomics data was analyzed using probabilistic graphical models, allowing functional characterization. Comparisons between groups were done using linear mixed models. Three proteomics tumor subtypes were defined. T1 (32% of patients) was related to adhesion, T2 (34%) had metabolic features, and T3 (34%) presented high splicing and nucleoplasm activity. These proteomics subtypes were validated in the PDAC TCGA cohort. Relevant biological processes related to carcinogenesis and tumor progression were studied in each subtype. Carcinogenesis in T1 subtype seems to be related to an increase of adhesion and complement activation nodes activity, whereas tumor progression seems to be related to nucleoplasm and translation nodes. Regarding T2 subtype, it seems that metabolism and, especially, mitochondria act as the motor of cancer development. T3 analyses point out that nucleoplasm, mitochondria and metabolism, and extracellular matrix nodes could be involved in T3 tumors carcinogenesis. Identified processes were different among proteomics subtypes, suggesting that the molecular motor of the disease is different in each subtype. These differences can have implications in the development of future tailored therapeutic approaches for each PDAC proteomics subtype.

## Introduction

Pancreatic adenocarcinoma (PDAC) is an aggressive disease with an overall 5 year-survival rate of only 5%. At the time of diagnosis, 80% of the tumors are already in incurable stages. On the other hand, in patients with localized disease, surgery represents the only possible curative treatment. However, despite performing a radical resection, 80% of the patients are going to relapse (1, 2). 60,430 new pancreatic adenocarcinoma cases and 48,220 related deaths have been estimated to occur in 2021 in the United States with an increasing incidence, being the fourth cause of cancer death (3). Therefore, it is an absolute priority deepen into the knowledge of pancreatic adenocarcinoma pathogenesis.

PDAC molecular subtypes have been already defined using transcriptomics data (4-6). Collison et al. divided PDAC into a classical, an exocrine-like, and a quasi-mesenchymal subtype (5). Moffit et al. established a classification making distinctions between tumor subtypes -basal-like and classical-, and stromal subtypes -normal and activated- (4). Finally, Bailey et al. divided PDAC into squamous, pancreatic progenitor, immunogenic and aberrantly differentiated endocrine exocrine (ADEX) subtypes (6). Squamous, quasi-mesenchymal and basal-like subtypes are pretty well aligned across the three classifications. Puleo et al. suggested that the differences showed by Bailey et al. were due to the cellularity of the samples (7).

Proteomics has been developed as a complementary approach to the massive sequence of genes and genomes and analysis at the RNA level. Its importance lies in the fact that proteins ultimately define the function and the operations of cells, tissues and organisms (8). Whereas genomics usually shows why things happen, proteomics explains what is happening. In this regard, genomics and proteomics complement each other integrating different levels of information.

A previous study defined proteomics subtypes of PDAC using hepatic metastases, classifying tumors into metabolic, progenitor-like, proliferative and inflammatory subtypes (9). Another study identified using proteomics four risk subgroups of PDAC (10). Recently, Cao et al. studied early biomarkers of PDAC by proteogenomics using tumor and normal adjacent tissues (11). However, until now, a proteomics study in order to define those processes involved in tumor development and progression has not been performed.

In this study, a molecular characterization of paired pancreatic adenocarcinoma samples (normal tissue-preneoplastic lesions-primary tumor-lymph node metastases) based on a proteomics analysis pipeline followed by computational approaches were performed to deepen the molecular information.

Coupling proteomics with our data analysis pipeline allow identifying those biological processes related to carcinogenesis and tumor progression through the analysis of paired samples. Network analysis based on probabilistic graphical models (PGMs) was used to further characterize those biological functions that may be relevant to tumor development and progression, comparing the different type of samples. Three proteomics tumor PDAC subtypes were identified and biological processes involved in carcinogenesis and tumor progression were different among them.

## Material & Methods

### Patient samples and clinical data

Patients with PDAC treated with surgery and adjuvant therapy from February 2010 to October 2020 at Hospital Universitario La Paz with available FFPE primary tumor and normal tissue, preneoplastic lesions grade 2-3 (PanIN), and/or lymph node metastases, were selected for the study. Samples were punched in order to study the differences associated with the different types of regions. A total of 52 primary tumors, 47 non-tumor tissues, 43 PanIN, and 31 lymph nodes were obtained for the proteomics analyses. The study was approved by the Ethical Committee from Hospital Universitario La Paz (IRB number: 1349).

### Protein isolation

Protein isolation was done as previously described (12). Briefly, FFPE sections were deparaffinized in xylene and washed twice in absolute ethanol. Protein isolates were prepared in 2% of SDS. Protein quantity was measured using MicroBCA Protein Assay Kit (Pierce-Thermo Scientific). Finally, 10 µg of each protein extract were digested with trypsin (1:50) and SDS was eliminated from the lysates using Detergent Removal Spin Columns (Pierce). Before mass-spectrometry experiments, samples were desalted using ZipTips (Millipore), dried, and resolubilized in 15 µL of a 0.1% formic acid and 3% acetonitrile solution.

Peptides were acidified to perform a stage-tip cleanup using two Empore reversed-phase extraction disks (3M) (13). Digests were dried in a SpeedVac and stored at &#x2212;20°C until LC-MS/MS analysis. Peptides were re-solubilized in 20 µl of 3% acetonitrile, 0.1% formic acid and 1 µl of indexed retention time (iRT)-peptides (Biognosys) were spiked in each sample for MS analysis. For the DDA analysis and subsequent spectral library generation, a small volume of each sample was taken and combined into a total of 10 pooled samples.

### Liquid chromatography-mass spectrometry experiments

One hundred and seventy-three samples from 52 PDAC patients including non-tumor tissue, primary tumor, PanIN and affected lymph nodes were analyzed by high-throughput proteomics.

Mass spectrometry analysis was performed on an Orbitrap Fusion (Thermo Scientific) equipped with a Digital PicoView source (New Objective) and coupled to a M-Class UPLC (Waters). Solvent composition of the two channels was 0.1% formic acid for channel A and 0.1% formic acid, 99.9% acetonitrile for channel B. For each sample 2 μl of peptides were loaded on a commercial MZ Symmetry C18 Trap Column (100Å, 5 µm, 180 µm x 20 mm, Waters) followed by nanoEase MZ C18 HSS T3 Column (100Å, 1.8 µm, 75 µm x 250 mm, Waters). The peptides were eluted at a flow rate of 300 nl/min. After an initial hold at 5% B for 3 min, a gradient from 5 to 22% B in 109 min and 32% B in 8 min was applied. The column was washed with 95% B for 5 min and afterwards the column was re-equilibrated to starting conditions for additional 10 min.

For library generation using the pooled samples, the mass spectrometer was operated in data-dependent mode (DDA) acquiring a full-scan MS spectra (350&#x2212;1’500 m/z) at a resolution of 120’000 at 200 m/z after accumulation to a target value of 400’000. Data-dependent MS/MS were recorded in the Orbitrap using quadrupole isolation with a window of 1.4 Da and HCD fragmentation with 30% normalized collision energy (NCE). Orbitrap resolution was set to 30’000, maximum injection time to 54 ms with a target value of 50’000, and the cycle time was set to 3 s. Charge state screening was enabled. Singly, unassigned, and charge states higher than seven were rejected. Precursor masses previously selected for MS/MS measurement were excluded from further selection for 25 s, and the exclusion window was set at 10 ppm.

For the analysis of the individual samples, the mass spectrometer was operated in data-independent mode (DIA). DIA scans covered a range from 400 to 1100 m/z in windows of 20 m/z. The resolution of the DIA windows was set to 30000, with an AGC target value of 50’000, the maximum injection time set to Dynamic and a NCE of 30. Each instrument cycle was completed by a full MS scan monitoring 350 to 2000 m/z at a resolution of 120000.

The samples were acquired using internal lock mass calibration on m/z 371.1010 and 445.1200. The mass spectrometry proteomics data were handled using the local laboratory information management system (LIMS) (14) and all relevant data have been deposited to the ProteomeXchange Consortium via the PRIDE (http://www.ebi.ac.uk/pride) partner repository with the data set identifier PXD032076.

### Spectral library generation and protein quantification

A hybrid spectral library was generated using the Pulsar search engine and spectral library generation functionality in Spectronaut (14.0.200601.47784, Biognosys) applying the default parameter settings to DDA and DIA runs. Spectra were searched against a canonical SwissProt database for human and common protein contaminants (NCBI taxonomy ID9606, release date 20190709). Carbamidomethylation of cysteine was set as fixed modification, while methionine oxidation and N-terminal protein acetylation were set as variable modifications. Enzyme specificity was set to trypsin/P allowing a minimal peptide length of 7 amino acids and a maximum of two missed-cleavages. Precursor and fragment tolerance was set to Dynamic, respectively for the initial search. The maximum false discovery rate (FDR) was set to 0.01 for peptides and 0.01 for proteins. Protein quantification was performed in Spectronaut using the default settings. The quantitative data were extracted using the BGS Factory Report (default) and used for follow-up analyses. Stringent filtering of the extracted feature groups by the Spectronaut reported q-Value was applied. For precursor fragment groups we required a per run q-value of at most 0.05 and a per experiment q-value of at most 0.01. The q-value sparse mode was used in combination with a global imputing strategy. To perform statistical modeling, fragment intensities were aggregated into precursor and peptide intensities.

### Data preprocessing

Proteomics data was transformed into log2. At least 75% of valid values in at least one group (non-tumor tissue, PanIN, primary tumor, and lymph nodes) was applied as quality criterion. Then, missing values were imputed to a normal distribution using Perseus software (15).

### Study of GATA6 expression by immunohistochemistry

For GATA6 determination, optimal tissue blocks were selected by an expert pathologist on haematoxylin and eosin (H&E) slides. Representative tumor areas of each case were selected for tissue microarray (TMA) construction. Two representative cores of 1.2 mm in diameter were taken and arrayed into a receptor block using a tissue microarrays (TMA) workstation (Beecher Instruments, Silver Spring, MD, USA) as previously described (16). 4 µm sections of the TMAs were used for immunohistochemistry (IHC) purposes. Briefly, slides were cut with a semiautomatic microtome HM 3508 (MICROM), deparaffinized and rehydrated in water. Antigen retrieval was performed in a DAKO PT Link. Peroxidase activity was blocked with Dako Protein block for 10 minutes, incubated for 30 minutes with primary antibodies, detected with Dako Envision Plus kit, and counterstained with haematoxylin. All reagents are from Dako (Agilent, CA, USA). GATA-6 antibody used: ref. n° AF1700 (R&D Systems, MN, USA).

### Probabilistic graphical models

As in previous works (12, 17), probabilistic graphical models (PGMs) were calculated using proteomics data without any a priori information in R using **grapHD** package (18). This analysis allows organizing protein data according their expression profile and identify relevant biological processes. The resulting networks were split into functional nodes accordingly to the gene ontology of its branches. Gene ontology analyses to assign a function to each functional node were done in DAVID webtool (19), using homo sapiens as background and GOTERM-FAT, Biocarta and KEGG as categories. Once the functional nodes were assigned, functional node activities were calculated as the mean expression of those proteins involved in the main function of each node. These functional node activities were used to make comparisons between groups of samples.

### Statistical analyses

Hierarchical cluster (HCL) based on correlation and average linkage using to establish tumor proteomics subtypes, were done using MeV software (20). Mixed linear models with fixed effects were used to establish the significant differences between groups of samples. These calculations were done in R using the library lme4 (21). For the comparison between tumor samples, a Mann-Whitney test was used. Finally, the relationships between clinical parameters and subtypes were studied using Chi-squared tests. These tests were done using Graph Pad Prism v6; p-values were two-sided and considered significant below 0.05.

### Validation of PDAC proteomics subtypes in TCGA cohort

A centroid based on the 313 differential proteins defined in the SAM was calculated for each tumor proteomics subtype. On 284 of these 313 proteins an equivalent gene existed in the TCGA cohort. Using these 284 genes, TCGA samples were classified in one of the three defined subtypes. Then, functional node activities were calculated to verify that the subgroups had the same molecular features than the tumor proteomics subtypes.

## Results

### Clinical data

From a cohort of 110 PDAC patients treated with surgery and adjuvant therapy from February 2010 to October 2020 at Hospital Universitario La Paz, fifty-two patients were selected for proteomics experiments. For these patients all the available samples were analyzed: non-tumor tissue, preneoplastic lesions grade 2-3 (PanIN), tumor, and lymph nodes. 47 non-tumor samples, 43 PanIN, and 31 lymph node samples were available.

Regarding clinical data, only information about fifty patients was available due to loss of follow-up after surgery of two of them. These fifty patients were used for the analyses that involved clinical parameters (Table 1).

**Table 1:** Patients’ characteristics.

| Number of patients = 50 (100%) |  |  |
| --- | --- | --- |
| Gender |  |  |
|  | Male | 30 (60%) |
|  | Female | 20 (40%) |
| Age (mean) |  | 28-84 (63) |
| Diabetes at diagnosis |  |  |
|  | Yes | 9 (18%) |
|  | No | 40 (80%) |
|  | Unknown | 1 (2%) |
| Tobacco use |  |  |
|  | Yes | 22 (44%) |
|  | No | 21 (42%) |
|  | Unknown | 7 (14%) |
| Location of primary tumor |  |  |
|  | Head | 38 (76%) |
|  | Body | 3 (6%) |
|  | Tail | 5 (10%) |
|  | Various | 4 (8%) |
| Grade |  |  |
|  | Very differentiated | 6 (12%) |
|  | Moderately | 33 (66%) |
|  | Poor | 8 (16%) |
|  | Unknown | 3 (6%) |
| Type of resection |  |  |
|  | R0 | 16 (32%) |
|  | R1 | 34 (68%) |
| pT |  |  |
|  | 1 | 3 (6%) |
|  | 2 | 11 (22%) |
|  | 3 | 34 (68%) |
|  | 4 | 2 (4%) |
| pN |  |  |
|  | N0 | 11 (22%) |
|  | N1 | 39 (78%) |
| Stage (TNM) |  |  |
|  | Ia | 2 (4%) |
|  | Ib | 2 (4%) |
|  | IIa | 4 (8%) |
|  | IIb | 38 (76%) |
|  | III | 3 (6%) |
|  | IV | 1 (2%) |

The median of follow-up was 13 months and 37 relapses had occurred, of which 10 were local relapses and 27 were distant relapses. All patients were treated with surgery and adjuvant therapy, and none of them received neoadjuvant therapy.

### Proteomics experiments

3,927 proteins were identified in DIA mass-spectrometry experiments. After applying a quality criterion of at least 75% of valid values in at least one group (non-tumor tissue, PanIN, primary tumor, and lymph nodes), 2311 proteins were used for the subsequent analyses.

### Proteomics pancreatic ductal adenocarcinoma subtypes

At first, all the samples were analyzed by a hierarchical cluster (HCL) to establish differences between different types of tissues. Surprisingly, the HCL was not capable to split samples by tissue type, i.e., establishing a group of non-tumor tissue, another of tumor samples, another with PanIN and a last one containing the lymph node samples (S1 Fig).

In order to establish if the variability associated with this distribution that does not distinguish by sample origin was related to different proteomics tumor subtypes, only tumor samples were selected to perform the analysis. In this case, the HCL clearly established three different groups of tumors in PDAC according to their proteomics profile. T1 included 16 (32%) patients whereas T2 and T3 was composed by 17 (34%) patients each one (Fig 1).

**Figure 1:**
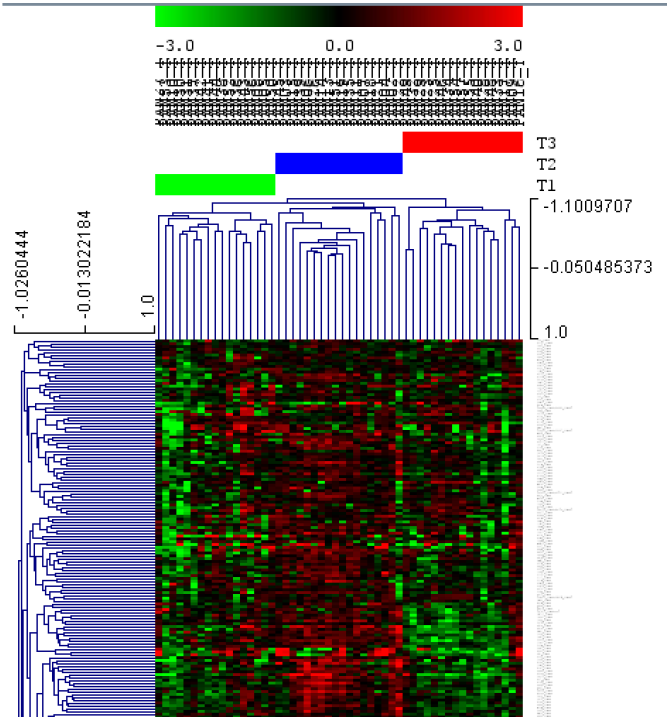
Hierarchical clustering (HCL) of PDAC tumor samples clearly showed three proteomics subtypes (T1, T2, and T3). HCL is based on average linkage method and Pearson correlation.

As in previous works (12, 17), a network analysis based on PGMs were used to characterize in depth the differences at biological processes level between the three proteomics PDAC tumor subtypes. The resulting network was divided into eight functional nodes, two of them with an overrepresentation of adhesion proteins. Functional node activities showed differences between the three subtypes. T1 presented higher functional node activities in adhesion and complement activation nodes, and will be referred as “adhesion subtype” for now on. T2 had higher functional node activities of mitochondria and metabolism and translation nodes, being for now on, the “metabolic subtype”. Finally, T3 showed higher functional node activities in nucleoplasm and splicing, and will be named as “nucleoplasm subtype” (Fig 2).

**Fig 2:**
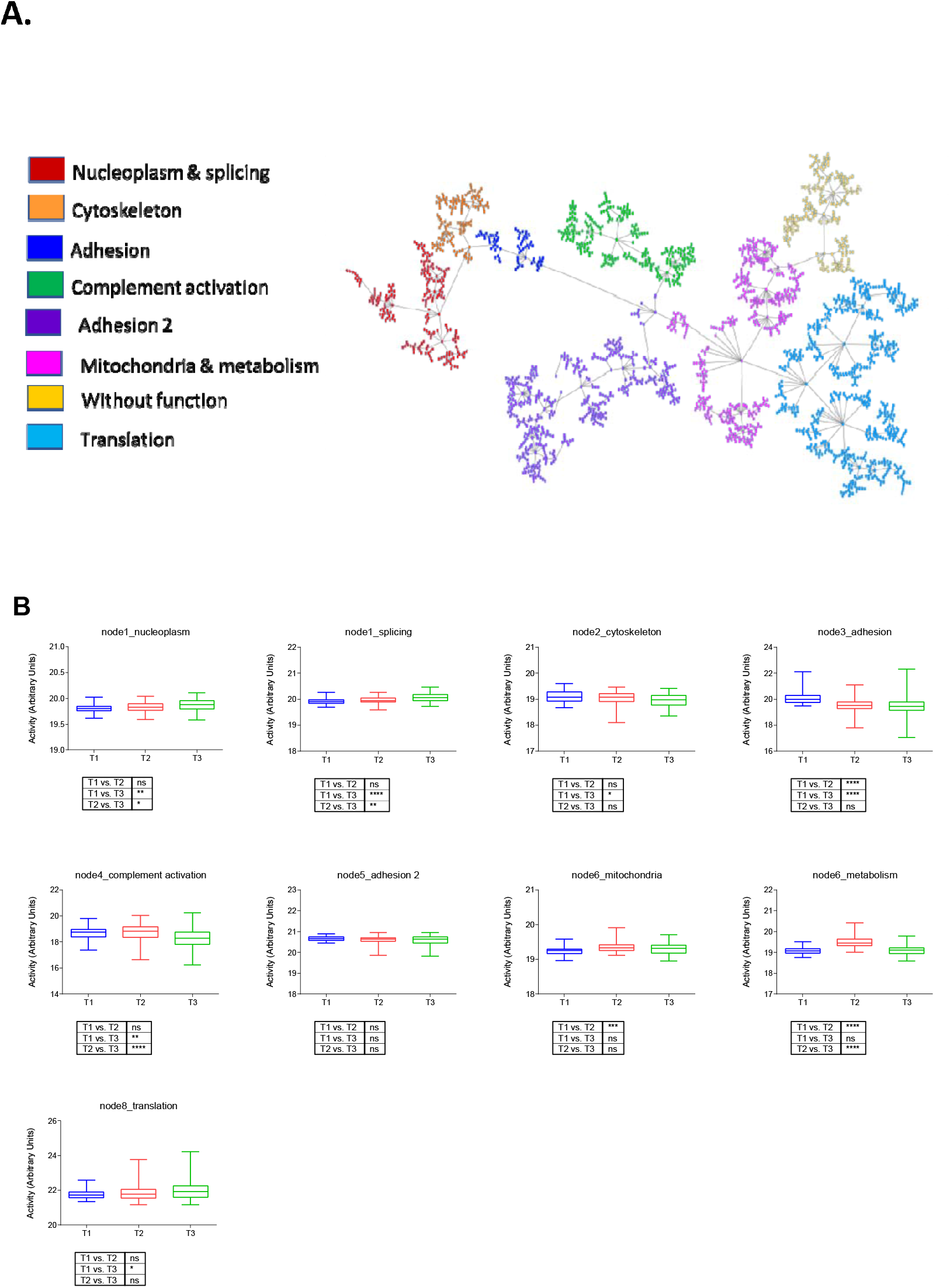
A. Network formed by 2311 proteins in PDAC tumor samples. B. Functional node activities comparing the three proteomics subtypes in tumor samples. ****: p<0.0001; ***: 0.0001<p<0.001; **: 0.001<p<0.05.

Regarding the clinical relevance of these subtypes, T1 and T2 contained most of the pancreatic tumors located in the head of the pancreas and T3 contained most of the tumors located in the body and tail (Sup Figure 1A). There were not significant differences between T groups according to gender, diabetes, pancreatitis, smoking, grade, type of resection, pT, pN, stage or location of metastases. Any differences in prognosis according overall survival or disease-free survival between the three PDAC proteomics subtypes were founded (Sup Fig 1B). The percentage of relapses at 12 months was 37% in T1 subtype, and 53% in T2 and T3 subtypes.

### Study of classical defined biomarkers from PDAC transcriptomics subtypes

Of the defined biomarkers from transcriptomics PDAC subtypes, only Mucin 5 (MUC5A), characteristic of Moffit classical subtype and Bailey progenitor subtype, and insulin (INS), characteristic of Bailey’s ADEX subtype, were identified in the list of the identified and quantified proteins. MUC5A expression was compared across the defined PDAC proteomics subtypes and it was significantly higher in T1-adhesion subtype. Additionally, insulin protein (INS) had a higher expression in T2 subtype, being comparable with ADEX subtype (Sup Fig 2).

GATA6, a marker characteristically expressed in Moffit classical subtype, was studied by IHQ. All T3 tumors showed a positive expression of GATA6 by IHC, and negative ones were split into T1 and T2 subtype (Sup Fig 3). Altogether, these results suggested that T2 tumors correspond to ADEX subtype and contained classical and basal-like tumors; T1 also contained basal-like and classical subtypes; and T3 corresponded only to classical tumors that also had an overexpression of proteins related to nucleoplasm.

**Fig 3:**
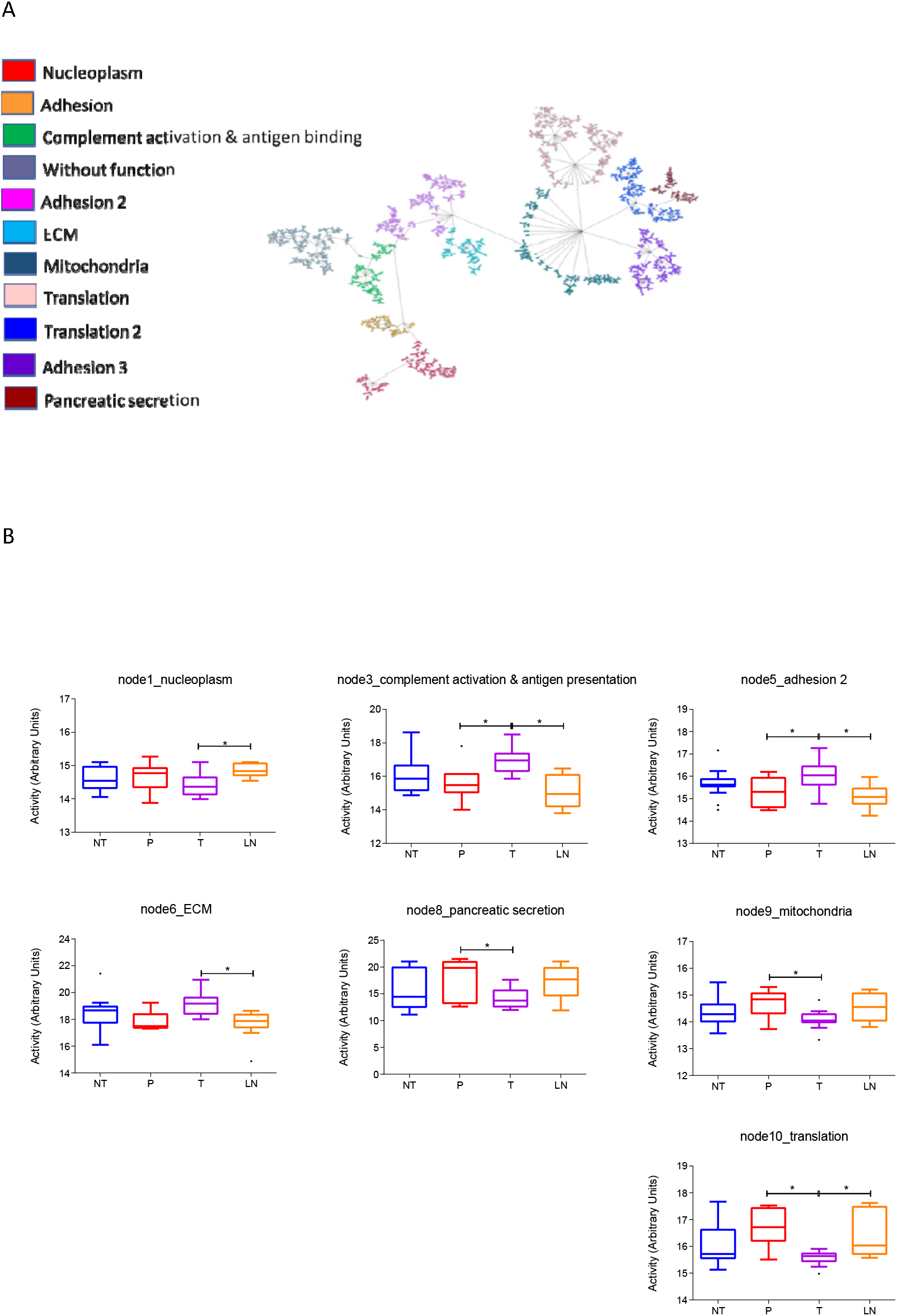
A. Network of 2311 proteins in T1 subtype. B. Differential functional node activities comparing the different histological samples in T1 subtype according to mixed linear models. NT= normal tissue, P= preneoplastic lesions, T= primary tumors, LN= lymph nodes. ****: p<0.0001; ***: 0.0001<p<0.001; **: 0.001<p<0.05; *: p<0.05

### Identification of biological processes involved in carcinogenesis and tumor progression in each PDAC proteomics subtype

Since differences between subtypes are bigger than differences between the types of the samples, we studied the differences between samples at a functional level in order to characterize the biological processes involved in the progression of the disease independently in each defined proteomics subtype. Thus, new analyses including each type of sample (normal pancreatic tissue, PanIN, tumor and lymph nodes) were performed for each proteomics subtype.

#### 1. Identification of T1 carcinogenesis and tumor progression processes

T1 tumor samples are characterized by a higher adhesion and complement functional nodes activities comparing with the other PDAC proteomics tumor subtypes.

A network based on PGMs was constructed including all types of samples from patients with T1 tumors. The resulting network had eleven functional nodes, one without an overrepresented biological function (Fig 3A). Functional node activities and mixed linear models were used to define those biological processes with differential functional node activities between tissue samples (Fig 3B, Table 2).

**Table 2:** Summary of functional node activities identified as differential using mixed linear models between samples in each PDAC proteomics subtype. NT= no tumor tissue, P= preneoplastic lesions, T= primary tumor, LN= lymph node metastasis.

| Samples | Direction | T1 | T2 | T3 |
| --- | --- | --- | --- | --- |
| NT→T | ↓ |  |  | ECM |
|  | ↑ |  | Pancreatic secretion<br>Metabolism | Nucleoplasm |
| P→T | ↓ | Pancreatic secretion<br>Mitochondria<br>Translation |  | Mitochondria &<br>metabolism |
|  | ↑ | Complement activation<br>Adhesion 2 | Pancreatic secretion<br>Metabolism |  |
| T→LN | ↓ | Complement activation<br>Adhesion 2<br>ECM | Pancreatic secretion<br>Metabolism |  |
|  | ↑ | Nucleoplasm<br>Translation |  |  |

Using mixed linear models, differences between non-tumor and tumor tissue were identified. Mitochondria, pancreatic secretion, and translation nodes activity decreased in tumor samples comparing to PanIN, and adhesion2, and complement activation and antigen presentation nodes activity presented an increase in tumor samples comparing to PanIn.

Significant differences between tumors and lymph nodes and therefore related to tumor progression were identified in complement activation and antigen presentation, adhesion 2, ECM, nucleoplasm, and translation functional nodes. In this case, nucleoplasm and translation were higher in lymph nodes and the others suffered a decrease in their activity in lymph node samples.

Adhesion 2 node contains some relevant proteins such as HSPB1 or THY. Complement activation node was mainly formed by immunoglobulins and complement proteins as C3 or C1QB. The translation node was mainly formed by ribosomal proteins (RPL3, RL23A, RL11, RL8, RS9, etc.). Finally, the most relevant protein included in nucleoplasm node was HIF1AN.

#### 2. Identification of T2 carcinogenesis and tumor progression processes

T2 tumors were characterized as a higher mitochondria, metabolism and translation activity comparing to the other PDAC tumor subtypes and presented overlapping characteristics with ADEX subtype. Again, a network was built using all T2 samples. It was composed of 12 functional nodes, two of them with two associated functions: pancreatic secretion and metabolism, and the other cytoskeleton and MAPK (Fig 4A).

**Fig 4:**
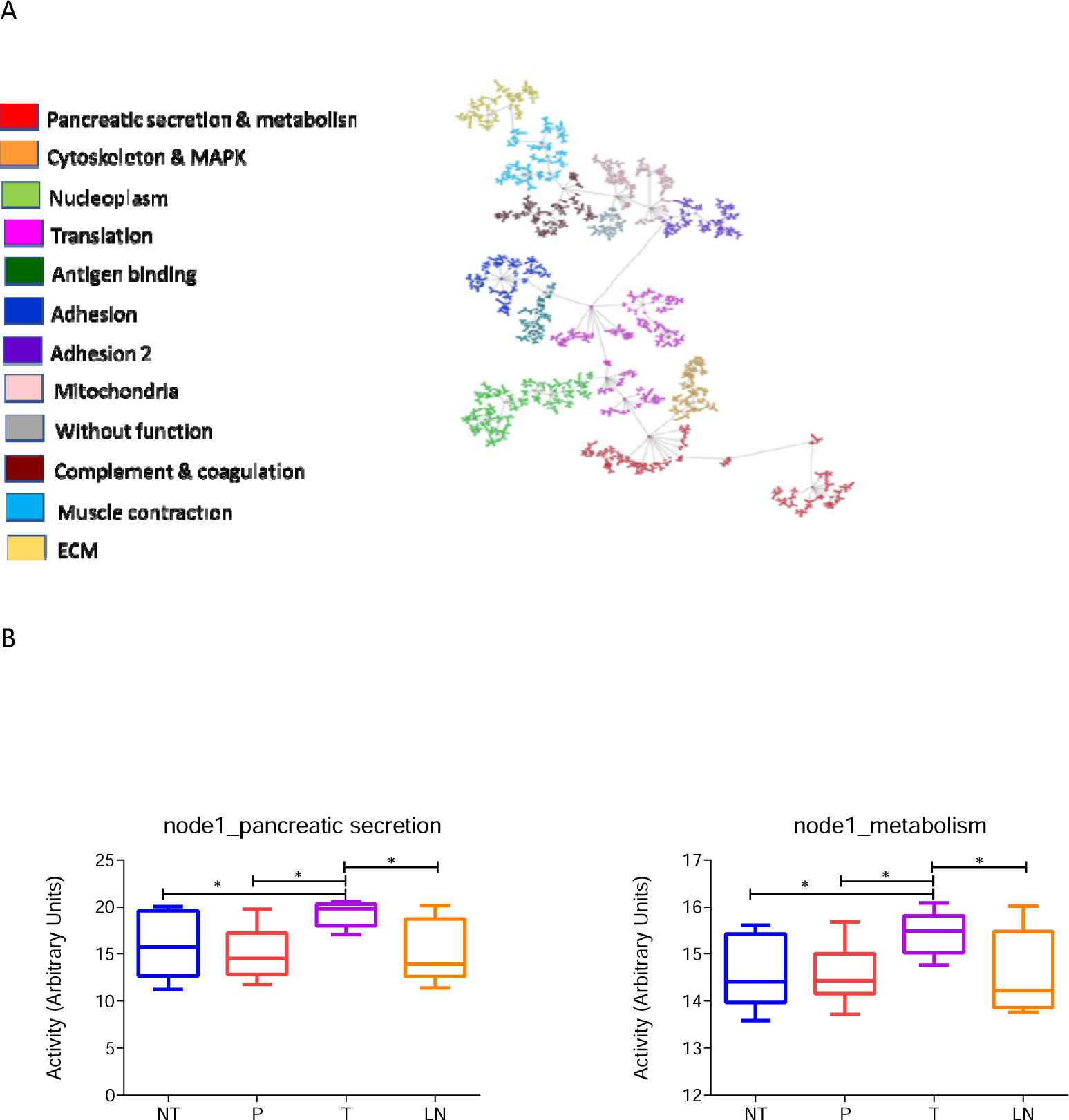
A. Network of 2311 proteins in T2 subtype. B. Differential functional node activities comparing the different histological samples in T2 subtype according mixed lineal models. NT= normal tissue, P= preneoplastic lesions, T= primary tumors, LN= lymph nodes. ****: p<0.0001; ***: 0.0001<p<0.001; **: 0.001<p<0.05; *: p<0.05 A

Functional node activities and mixed linear models were used to define those biological processes with differential functional node activity between tissue samples (Fig 4B).

Those biological processes identified as related to tumor development were pancreatic secretion and metabolism, which were significantly higher in tumor samples than in normal tissues and PanIN.

These functional node activities (pancreatic secretion and metabolism) presented a significant decrease in lymph node samples comparing to tumors, being able to be associated with tumor progression.

Pancreatic secretion node contained several relevant proteins such as TYMP, NAMPT or pancreatic lipases as PNLIP.

#### 3. Identification of T3 carcinogenesis and tumor progression processes

T3 subtype was characterized by a higher nucleoplasm activity and also had overlapping characteristics with classical subtype. The obtained network using protein expression data from T3 samples was composed by ten functional nodes (Fig 5A).

**Fig 5:**
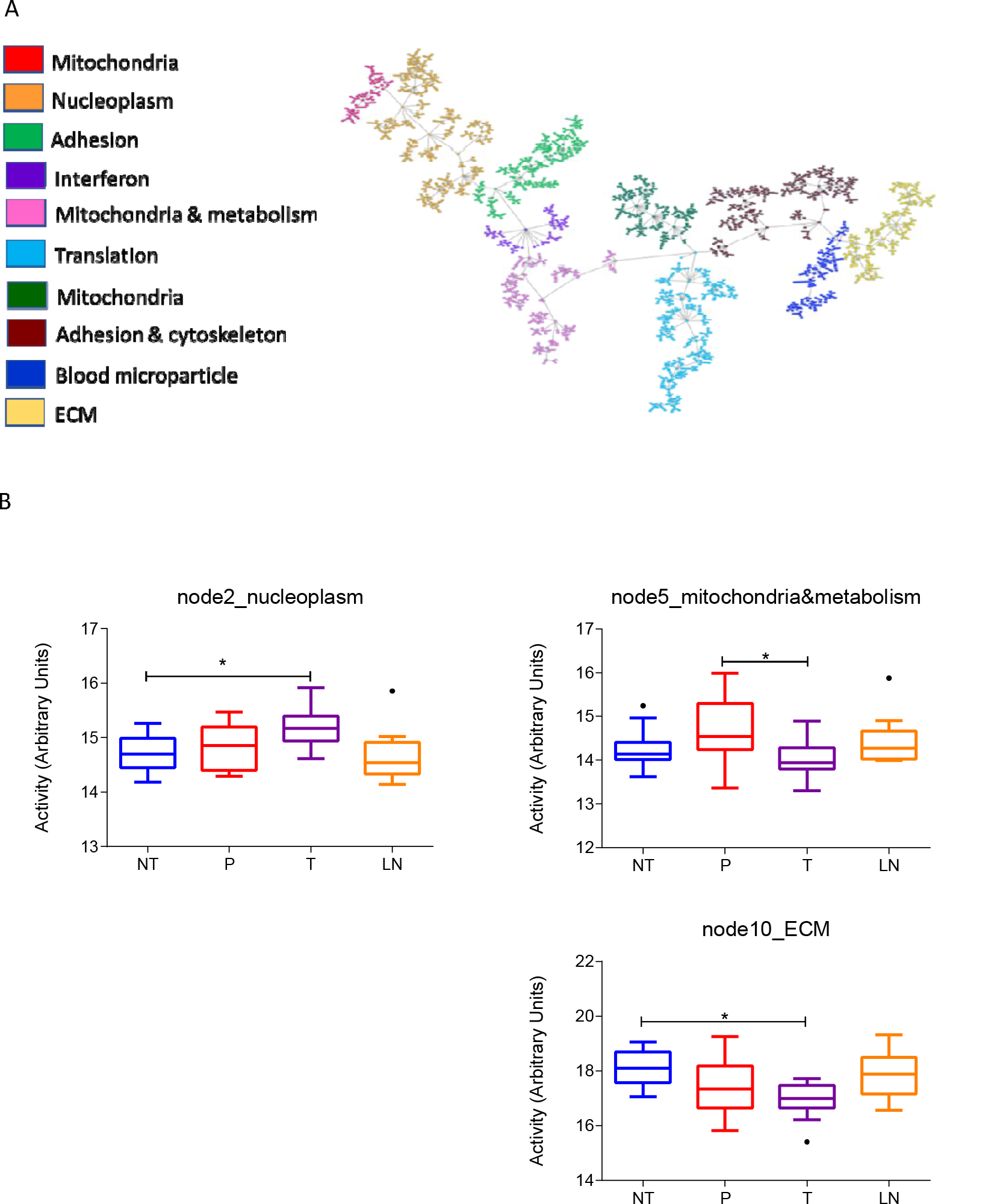
A. Network of 2311 proteins in T3 subtype. B. Differential functional node activities comparing the different histological samples in T3 subtype according mixed lineal models. NT= normal tissue, P= preneoplastic lesions, T= primary tumors, LN= lymph nodes. ****: p<0.0001; ***: 0.0001<p<0.001; **: 0.001<p<0.05; *: p<0.05

Functional node activities showed differences between no tumor and tumor samples in nucleoplasm, mitochondria & metabolism, and ECM. ECM had a decrease in their activity in tumors comparing to normal samples while nucleoplasm showed an increase in tumor samples. In addition, tumor samples presented a decrease in mitochondria & metabolism node activity compared to PanIN. In the case of tumor and lymph nodes, there are not any processes significantly different between them (Fig 5B).

Nucleoplasm node was formed by some well-known proteins as PARP1, ELAVL1, SART3, RAN, FUBP1, APEX1 or AKT1S1.

A summary of the differential functional node activities and its corresponding biological processes is presented in Table 2. Complete results of mixed linear models are provided in Sup File 1.

### Validation of these tumor subtypes in the TCGA cohort

In order to confirm the described PDAC proteomics subtypes, the TCGA cohort was used. According to a centroid assignation, there were 46 (25%) PDAC samples in T1 subtype, 73 (40%) in T2, and 65 (35%) samples in T3 subtype. Using functional node activities calculated in this cohort confirmed that the TCGA samples assigned to T2 subtype had metabolic characteristics. T1 subtype samples had higher activities in adhesion node as it occurred in the proteomics cohort. T3 subtype showed a higher activity in nucleoplasm and splicing functional node (Sup Fig 4). None differences in overall survival between the tumor proteomics subtypes were found.

## Discussion

This is the first study in PDAC using proteomics to define molecular subtypes and mechanisms involved in tumor development and progression in each subtype. Samples from 52 PDAC patients including non-tumor tissues, preneoplastic lesions, primary tumors and lymph nodes, were analyzed by high-throughput proteomics and a Systems Biology approach in order to identify relevant biological processes in tumor development and progression.

Using this approach, we have defined three proteomics PDAC subtypes, which can be detected even in the earliest stages of tumor development. Each defined subtype showed specific molecular features. T1 subtype is related with adhesion, T2 subtype has metabolic features, and T3 subtype presented high splicing and nucleoplasm activity. These proteomics subtypes also shared some characteristics with subtypes previously defined by transcriptomics, while providing new and complementary information: T2 tumors correspond to ADEX subtype, including some metabolic basal-like and classical subtypes; T1 contained basal-like and classical subtypes; and T3 corresponded to those classical tumors with high expression of nucleoplasm related proteins. Interestingly, identified processes involved in tumor development and progression were different between the three PDAC proteomics subtypes, suggesting that the molecular motor of the disease is different in each subtype. These differences can have implications in the development of future tailored therapeutic approaches for each PDAC proteomics subtype.

Previous transcriptomics studies defined a group of tumors where adhesion plays an important role (4-6), as observed in our proteomics subtype T1. Carcinogenesis in T1 subtype seems to be related to a decrease of mitochondria, pancreatic secretion and translation nodes activity, and an increase of adhesion and complement activation and antigen presentation nodes activity. Adhesion 2 functional node contains some relevant proteins, such as HSPB1 and THY1. HSPB1 gene codifies Heat Shock Protein 27 (Hsp27), a cell survival protein found at elevated levels in many human cancers including prostate, lung, breast, ovarian, bladder, renal, pancreatic, multiple myeloma and liver (22, 23). THY1, also known as CD90, is a stem cell marker that interacts with monocytes and macrophages, promoting immunosuppressive features of immune cells, enhancing the stemness and E-MT of PDAC. It has been suggested that THY1 establishes a favorable environment that promotes tumor progression (24), which can be mediated by high levels of PD-L1 in CD90^+^ cells (25). In addition, complement and antigen activation functional node was mainly composed by immunoglobulins and complement proteins. Although the role of complement in PDAC development is still unclear (26), the role of complement in tumor development and modulation of tumor microenvironment has been demonstrated (27). The expression of complement C3 in pancreatic cancer was described as significantly higher than in normal tissues, being proposed as a diagnostic biomarker of early-stage pancreatic cancer (28, 29). Depletion of C3 in tumor cells enhanced efficacy of anti–PD-L1 treatment (30). These results together suggest that high levels of THY1 and complement components in T1 tumors provoke and immunosuppressive tumor microenvironment, suggesting an inflammatory phenotype, and open up the possibility of using a combination of immunotherapy coupled with anti-PD1/PD-L1 therapy in patients with these T1 tumors. Differences between tumors and lymph nodes in T1 subtype, related to tumor progression, were identified in nucleoplasm, translation, adhesion, extracellular matrix, and complement activation nodes. In nucleoplasm functional node HIF1AN stands out, due to its role in the regulation loop of IGFR. It has been described that the use of an IGFR inhibitor caused a lower expression of this protein and a decrease in growth in pancreatic cancer cells (31, 32) Thus, IGFR pathway inhibitors may avoid tumor progression in PDAC T1 proteomics subtype.

Our data in primary tumors confirmed that mitochondria metabolism plays an important role in one of the PDAC proteomics subtypes, T2 subtype. Our analysis based on probabilistic graphical models also highlighted the importance of glycolysis and pyruvate metabolism, valine metabolism, and fatty acid metabolism among others. In a previous study analyzing tumor and adjacent tissue from three PDAC patients, differential proteins related to metabolism, especially mitochondrial proteins and proteins whose function is acting as regulators of pancreatic juices, were identified (33). In addition, in a previous proteomics study in hepatic PDAC metastases, a group related to metabolism was defined, characterized by the expression of ethanol oxidation, mitochondrial fatty-acid beta oxidation and retinoic acid signaling pathways (9). Metabolism and pancreatic secretion nodes had differential activity between T2 normal tissue, PanIN and tumors, and also between tumors and lymph nodes. Pancreatic secretion node contained some relevant proteins. For instance, expression of the angiogenic factor TYMP has been correlated to capecitabine and fluorouracil response (34, 35). Additionally, Law et al. established in their proteomics study that TYMP had a strong correlation with patient survival in PDAC (9). Another protein in this node is NAMPT, whose inhibitor STF-118804, in combination with chemotherapy agents such as paclitaxel, gemcitabine, and etoposide, showed an additive effect in the decrease of cell viability and growth in PDAC (36). PNLIP is one of the main pancreatic lipases. It is related to orlistat, a drug used in obesity treatment. Kridel et al. state that orlistat may inhibit growth of prostate cancer by interfering with the metabolism of fats (37). Interestingly, in lymph node metastases these processes presented a significant decrease comparing to primary tumors. In conclusion, regarding T2 subtype, it seems that metabolism and, specially, mitochondria act as the motor of cancer development.

The last proteomics subtype, T3, was related to nucleoplasm and histones. Mutational studies of PDAC showed a high prevalence of genetic alterations in genes involved in chromatin remodeling such as *SMARCA2, SMARCA4, MLL2* or *ARID1A*, among others (38), so it is not surprising that proteomics subtyping highlighted the relevance of proteins related to nucleoplasm and histone modification. This group also was GATA6 positive, being equivalent to classical PDAC tumors. T3 analyses point out that nucleoplasm, mitochondria & metabolism, and extracellular matrix nodes could be involved in T3 tumors carcinogenesis. The nucleoplasm node also contained some well-known cancer-related proteins, such as PARP1. ELAVL1 is also present in this functional node and it has been associated with response to gemcitabine in pancreatic cancer (39). SART3 is an RNA-binding nuclear protein that is a tumor-rejection antigen. This antigen possesses tumor epitopes capable of inducing HLA-A24-restricted and tumor-specific cytotoxic T lymphocytes in cancer patients and may be useful for specific immunotherapy. RAN promotes metastasis and invasion in pancreatic cancer by deregulating the expression of AR and CXCR4. In this study they also demonstrated that the expression of Ran was remarkably higher in lymph lode metastases than in primary pancreatic cancer tissue (40). FUBP1 is a target of irofulven, a novel anti-cancer compound, whose anti-tumor activity in an advanced pancreatic cancer patient was documented (41). APEX1 redox selective inhibitor E3330 caused a significant inhibition of tumor cell migration in PDAC (42). AKT1S1 is a target for rapamycin, a drug used in the treatment of other cancers (43, 44).

Proteomics has been previously used to characterize PDAC disease employing serum, pancreatic juice, fresh tissue and paraffin samples. Holm et al. analyzed 21 serum samples from patients with pancreatic cancer to identify proteins differentially expressed between patients with long or short survival (45). Paulo et al. compared PanIN lesions and PDAC FFPE samples, identifying a list of exclusive proteins for each condition. Annexin 4A, fibronectin and mucin 2 were exclusively expressed in PDAC samples (46). Naidoo et al. conducted the first study of FFPE samples comparing PDAC and lymph node metastases and found that proteins differentially expressed were mostly related to immune system and metabolic processes (47). Cao et al. recently identified some proteins that could be useful as early detection biomarkers in PDAC comparing normal and tumor tissue (11). However, in these studies the differences between molecular subtypes were not evaluated. In this context, our approach has two main advantages: first, analyzing different stages of tumor progression (non-tumor tissue, PanIN, tumor and lymph nodes) allowed us to study carcinogenesis (differences between PanIN and tumor tissue) and tumor dissemination (differences between tumor tissue and lymph nodes) independently. Second, our analytical pipeline allows studying biological processes instead of proteins individually, providing naive and undirected context to the high-throughput proteomics data and allowing interpretation of the molecular features detected in each proteomics subtype. Additionally, our proteomics subtypes were validated by the PDAC TCGA cohort.

Remarkably, our analyses showed that differences between tumor subtypes are higher than between type of tissues. Connor et al. analyzed 19 paired samples, primary tumors and metastases, and showed that they were molecularly conserved, i.e., paired metastases and primary tumors were classified in the same molecular subtype (48). The fact that adjacent non-tumor tissue is more related to its neighbor tumor than non-tumor tissue from other patients suggests that the physical tumor border does not correspond with the molecular tumor border in pancreatic adenocarcinoma.

Drug development in PDAC is challenging, as modest results of immunotherapy in this pathology points out. Although several reasons for this lack of results have been proposed (49), the inclusion of unselected patients in clinical trials, regarding its molecular features, may be a hidden factor, point out the need of taking molecular heterogeneity of PDAC into account in future developments. Our results suggest some therapeutic strategies to follow up in each proteomics subtype. For instance, regarding T1 tumors, HSPB1 is target of the drug apatorsen, a second-generation antisense drug able to inhibit the production of Hsp27 in preclinical experiments. Data from the RAINIER trial showed adding apatorsen to gemcitabine+nab-paclitaxel did not improve the outcome of unselected metastatic PDAC patients, but can be useful in those patients with high serum doses of Hsp27 (50). Additionally, high levels of THY1/PD-L1 and complement components in T1 tumors provoke an immunosuppressive tumor microenvironment, suggesting an inflammatory phenotype (25), and depletion of C3 in tumor cells enhanced efficacy of anti–PD-L1 treatment (30). These results open up the possibility of using a combination of complement immunotherapy coupled with anti-PD1/PD-L1 therapy in patients with these T1 tumors. Regarding T2 tumors, mitochondria is emerging as an interesting actionable target, with numerous clinical trials currently testing different drugs modulating mitochondrial activity in PDAC (51). Finally, T3 tumors showed overexpression of a variety of actionable targets. Veliparib, a PARP-1/2 inhibitor, was tested with gemcitabine and radiotherapy in locally advanced pancreatic cancer in a phase 1 study and the results supported a phase 2 validation study (52).

The main limitation of this study was the impossibility to get all types of samples from each patient. This limitation was mitigated using linear mixed models. Additionally, after dividing samples by subtype, the number of samples in each group decreased, which may have prevented the detection of differences in the possible predisposing factors, clinical characteristics and prognosis of the different proteomic subtypes. In addition, all biological processes that might be therapeutic targets in the future needs further study

In this study, three PDAC proteomics subtypes were defined, an adhesion-related subtype (T1), a metabolic-related subtype (T2), and a nucleoplasm subtype (T3). We also suggested several biological processes involved in tumor development and progression characteristic of each proteomics subtype, suggesting that the motor of the disease is different in each subtype. These biological processes could be relevant as a guide to stratify patients and select candidates for future tailored therapeutics treatments in PDAC.

## Data Availability

All relevant data have been deposited to the ProteomeXchange Consortium via the PRIDE (http://www.ebi.ac.uk/pride) partner repository with the data set identifier PXD032076 and it will be available since the publication of the study.

## Author contributions

### Ethics Statements

This project has been approved by Ethics Committee of Hospital Universitario La Paz (PI-2043) and written consent was obtained for all the participants in the study.

