## Supplementary material for "Identification of carcinogenesis and tumor progression processes in pancreatic ductal adenocarcinoma using high-throughput proteomics": Sup fig

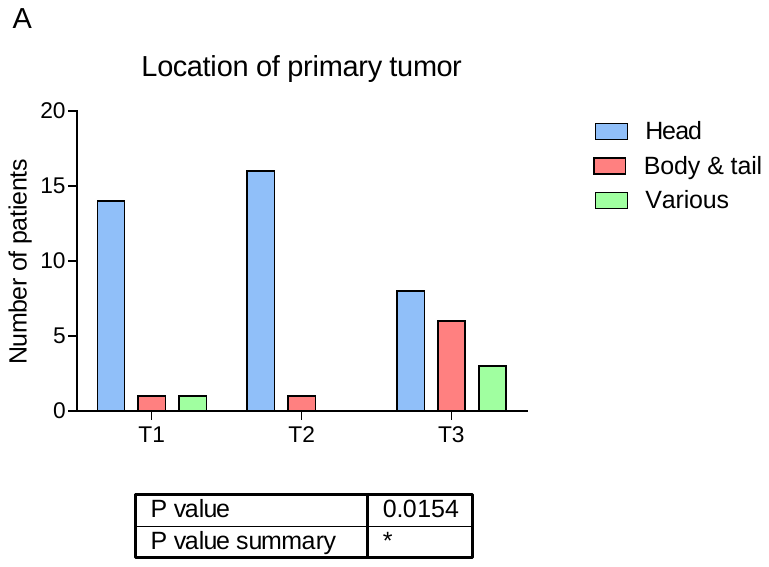

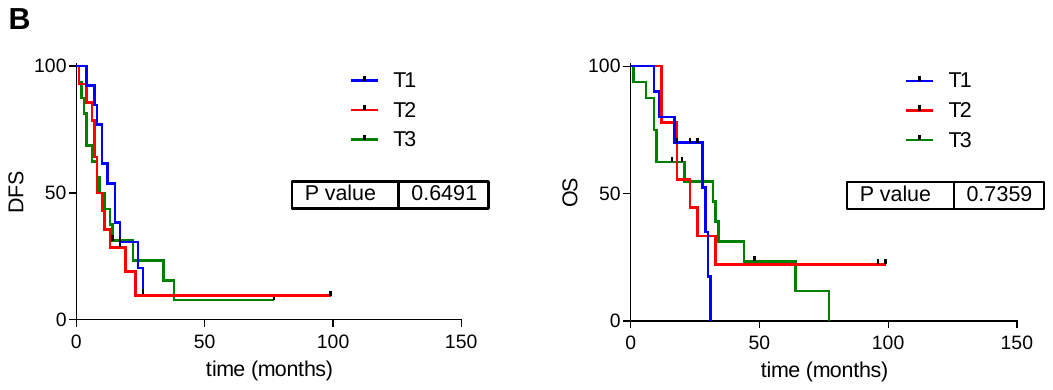

Sup Figure 1: A. Distribution according to the location of primary tumor in PDAC proteomics subtypes. B. Disease-free survival (DFS) and overall survival (OS) according to the three PDAC proteomics subtypes.

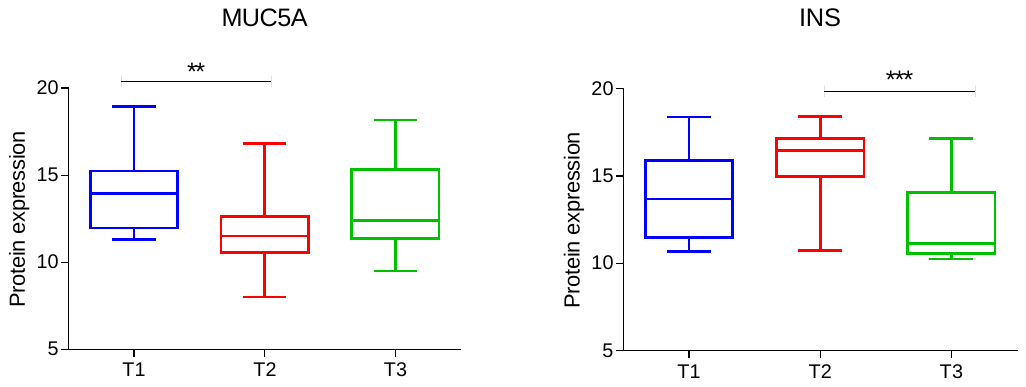

Sup Fig 2: Mucin-5 (MUC5A) and insulin (INS) expression in PDAC proteomics subtypes. ****: p<0.0001; ***: 0.0001<p<0.001; **: 0.001<p<0.05; *: p<0.05.

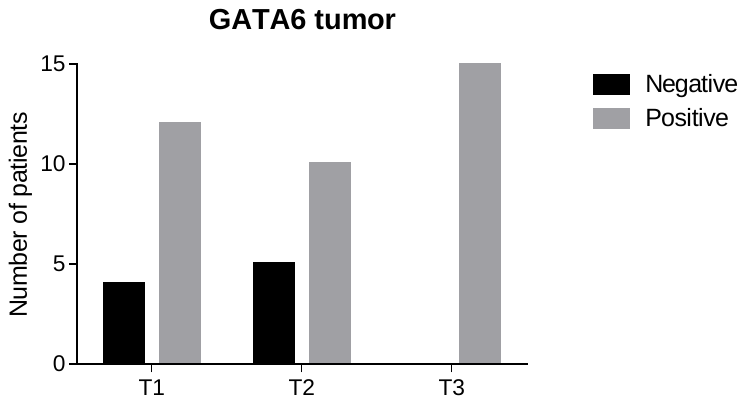

Sup Fig 3: Distribution of GATA6 immunohistochemical expression in PDAC proteomics subtypes.

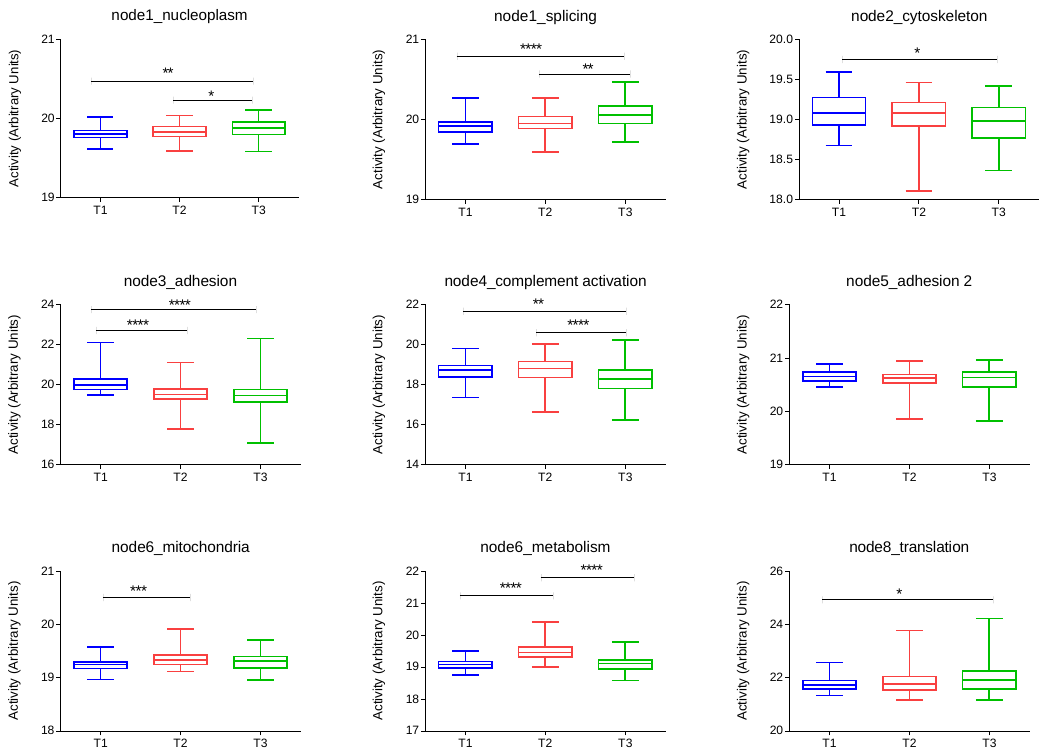

Sup Fig 4: Validation of the functional node activities in the TCGA cohort. ****: p<0.0001; ***: 0.0001<p<0.001; **: 0.001<p<0.05; *: p<0.05
